# Establishing the Effectiveness of Enhanced Implementation Methods in Preventative Behavioral Health

**DOI:** 10.1101/2020.09.25.20200444

**Authors:** Sarah A. Stoddard, Barrett Wallace Montgomery, Leah D. Maschino, Kristen Senters Young, Julia W. Felton, Amy Drahota, Jennifer E. Johnson, Debra Furr-Holden

## Abstract

**Background:** More than 20 million Americans ages 12 and older have a past-year substance use disorder. Majority-minority cities, including Flint, MI, suffer disproportionately from higher rates of substance use and are less likely to have access to evidence-based prevention and treatment interventions relative to predominately White and wealthier cities. Thus, identifying approaches that can improve implementation of evidence-based substance use practices is a critical public health goal. In the current report, we provide a detailed protocol for the implementation and evaluation of the Strengthening Flint Families initiative, a community-based implementation study of a multi-level behavioral health intervention that includes peer recovery coaching, the Strengthening Families Program, and a multimedia campaign. Our goal is to improve family resilience and reduce behavioral health disparities in the Flint community, as an example of how this could be done in other communities. Our overall strategy includes community-informed implementation enhancements to increase adoption and sustainment of evidence-based behavioral health services.

**Methods:** This project has 4 phases that align with study aims-1) a systematic assessment of behavioral health organizations in the Flint Area to understand organizational needs and strengths in Flint; 2) tailoring implementation strategies for one individual-level evidence-based practice, peer recovery coaching, and one family-level evidence-based program, the Strengthening Families Program; 3) building capacity and promoting sustainability; and 4) evaluating primary (implementation) and secondary (effectiveness) outcomes. Implementation outcomes are derived from the RE-AIM framework and effectiveness outcomes will be assessed at the individual, family, and community levels.

**Discussion:** Understanding and addressing the behavioral health organizational needs, strengths, and barriers to program adoption and referral in Flint offers great promise to strengthen the behavioral health network of providers serving Flint residents. Moreover, understanding barriers to accessing and implementing behavioral health services in low-resource communities may prove to be a valuable tool for discovering the most effective implementation methods tailored to specific organizations. These evidence informed approaches may prove useful for cities outside of Flint.

**Contributions to the literature:** More research is needed on how best to improve community capacity for substance use services, especially in low-income communities

This study will examine implementation strategies for improving community adoption and sustainment of a multilevel suite of substance use interventions

Study findings will contribute to improved community-wide implementation of evidence-based behavioral health services and increased access to and participation in these services in low-resource communities

## Background

In the United States, over 20 million Americans ages 12 and older (7.5% of the population) are classified as having a substance use disorder (SUD) in the past year (Substance Abuse and Mental Health Services Administration, 2017a). Poor, primarily minority cities, including Flint, Michigan, often face substance use problems that exceed the national average. In 2013, more than 15% of Flint residents participating in a community-wide survey on health reported that they or a family member used or were addicted to drugs in the past 2 years (Speak To Your Health, 2014). More recent local data have been collected but have not been made available to the public. Other indicators of substance use problems, such as opioid overdose death, have shown steady increases since 2013 (e.g., Sadler and Furr-Holden, 2019). Though substance use disorders are common, recurrent, and associated with excess morbidity and mortality, they are treatable, and people can and do recover.

Although multiple effective substance use interventions exist for adults and youth (e.g., Dugosh et al., 2016; MacArthur et al., 2016), barriers remain for individual utilization of these programs (Liebling et al., 2016), organizations to adopt programs (Barclay et al., 2019), and community-wide implementation of evidence-based services (Romero et al., 2017). The availability of substance use interventions does not ensure utilization of these programs by those in need (Wen, Druss, & Cummings, 2015; Wang et al., 2005; Green et al., 2009). Lack of access to and awareness of evidence-based services along with stigma are common barriers for utilization of substance use interventions (Liebling et al., 2016). At the organizational level, recruitment, efficient training of staff, and sustainability of implementing and funding programs are cited barriers to the success of substance use interventions (Barclay et al., 2019). Moreover, stigma and poor organizational collaboration can also act as barriers to successful implementation of evidence-based services (Romero et al., 2017).

To improve program uptake at the individual, organizational, and community-level it is important to understand and address the barriers to accessing and implementing evidence-based services while ensuring they are implemented with fidelity (Kilbourne et al., 2013). More research is needed to investigate these barriers, understand why they exist, and how they can be reduced to improve the utilization and implementation of evidence-based services. More specifically, while evidence-based services exist in Flint, Michigan, overdose deaths and substance misuse and abuse among Flint residents continue to increase (Sadler and Furr-Holden, 2019). The goal of the current project is to examine whether community-informed implementation strategies increase adoption, utilization, and sustainment of a multilevel suite of evidence-based substance use interventions. Improving the utilization and implementation of existing evidence-based services in Flint is hypothesized to reduce substance misuse and abuse rates.

## Methods / Design

Strengthening Flint Families (SFF) is a multi-level intervention designed to reduce behavioral health disparities and improve family resilience. SFF is a project of the Flint Center for Health Equity Solutions (FCHES) (Ellington et al., 2020). The Center has entered into a number of formal partnerships with organizations and citizens of Flint directly affected by, or helping to alleviate, substance use disorders in the community. SFF has also partnered with a local community-serving organization for this initiative. Our community partner organization, Flint Odyssey House, has decades of experience with substance use treatment and prevention in Flint. These collaborative efforts will explore and work to reduce barriers to the utilization and implementation of behavioral health evidence-based services in Flint, Michigan. Partnering with this organization and many others in Flint is an important part of Community-Based Participatory Research (CBPR) methods and principles (Israel et al., 2008). Green et al. defined CBPR as an “inquiry with the participation of those affected by an issue for the purpose of education and action for effecting change” (Green et al., 1995). This initiative has partnered with multiple community organizations, including behavioral health providers, the local health coalition, and governmental agencies on the design, execution, and evaluation of the initiative.

SFF includes the implementation of three effective behavioral health interventions to reduce behavioral health disparities and improve family resilience in the Flint Community. These interventions operate at the individual, family, and community levels, and together, are an excellent fit for the needs of economically distressed, minority-majority cities such as Flint. The interventions include; at the individual-level, Peer Recovery Coaches (PRCs); at the family-level, the Strengthening Families Program (SFP); and at the community-level: a multi-media campaign. While the SFP and PRC services are currently offered in Flint, this implementation research seeks to expand the reach and accessibility of these services as well as work to reduce barriers to access and successful community-wide implementation.

### Interventions

PRCs are certified professionals with lived experience of recovery and often work in substance use treatment centers. These recovery professionals provide a unique mentorship experience by helping to enroll community members in substance use treatment, supporting individuals during treatment, and assisting them as they step down levels of care (e.g., intensive outpatient to outpatient), transition out of treatment, or work to maintain their recovery. PRCs work one-on-one with clients in various settings throughout their recovery process. Development plans are created to guide the process and reach milestones along the way (Substance Abuse and Mental Health Services Administration, 2017b). PRCs and peer support have been shown to reduce the risk of relapse (Boisvert et al., 2008; Chinman et al., 2014) and inpatient admissions (Chinman et al., 2014).

The SFP is an evidence-based, family-level prevention program that has been effective in improving parenting skills and children’s mental health (depression, aggression, conduct disorders) in high-risk children of individuals receiving treatment for substance use (i.e., SFP 7-17; Kumpfer & DeMarsh, 1986; DeMarsh & Kumpfer, 1986) and in improving parenting skills and relationships, and reductions in delinquency and substance use in adolescents (i.e., SFP 10-14; Spoth et al., 2004; Kumpfer et al., 2002; Gottfredson et al., 2006). This study focuses on the model for families with children ages 10-14 (i.e., SFP 10-14; Spoth, Redmond, Shin, 1998). SFP 10-14 consists of seven sessions over the course of seven weeks. The program is typically taught to 5-10 families in a group format. Each group session begins with a family-style dinner. Following dinner, the youth and parents/caregivers meet separately to focus on youth well-being and parenting skills, respectively. Each session ends with a family session to review and practice the skills from the session.

Finally, at the community-level, a substance misuse prevention multi-media campaign will be developed and tailored specifically for Flint residents. The campaign will focus on messaging to reduce the stigma of recovery help-seeking and advertise the availability of services. The multi-media campaign will include a combination of modalities including social media (e.g., Facebook, Twitter, and Instagram), print media (e.g., bench advertisements in key neighborhoods), and radio content. The use of mass media to advertise service availability has been effective in promoting sustained behavior change across six different behavior domains, including addictive behaviors (Robinson et al., 2014).

### Study setting

This implementation study is being conducted in Flint, Michigan. The Flint community has many needs, including clear racial health disparities, entrenched problems with chronic disease, and deep economic distress (Sadler, Gilliland, & Arku, 2013; Sadler & Highsmith, 2016; Hanna-Attisha et al., 2015). The social and built environment characteristics of Flint, Michigan, make it a unique location for a study of mental health and substance abuse. Flint has experienced significant loss in both employment and population since the 1960s (Jacobs, 2009). Unemployment, abandonment of housing, and loss of services like police and fire have contributed to a huge increase in crime (Sadler et al., 2017), and other amenities like healthy food are extremely difficult to find (Shaver et al., 2018). Further, financial troubles brought on by poor urban policy at the state level contributed to the deterioration of the water network resulting in the Flint Water Crisis, which has differentially affected poor and minority residents in Genesee County (Hanna-Attisha et al., 2015; Sadler et al., 2017). Presently, 45% of Flint residents live below the Federal poverty line, and 57% of residents are African-American (US Census Bureau, 2017).

The Flint community also has a wealth of knowledge and experience generated by decades of community public health efforts and grassroots activism (Griffith et al., 2010). Therefore, it is ideally suited for a CBPR project that brings additional resources to ongoing community efforts and helps share what is learned with other communities. Over the years, community-based organizations in Flint have raised concerns over the focus on experimental research with no plan to sustain the benefits of results from previous research efforts, a sentiment which is in line with recent findings on the sustainability of evidence based interventions (Hailemariam et al, 2019). The lack of attention to dissemination and implementation research in Flint highlights the importance of an implementation-focused study (Bernet et al., 2013) and creating a self-sustaining public health program in this community.

### Aims of Strengthening Flint Families

This multi-level implementation study will primarily evaluate the ability of community-informed implementation strategies to promote reach, effectiveness, adoption, implementation, and maintenance of evidence-based individual, family, and community-level substance use interventions. Secondarily, we will monitor the effectiveness of these programs in low-income communities using the tailored implementation strategies. All study components were reviewed and approved by the Michigan State University Institutional Review Board (IRB# x17-1370e; STUDY00002533; STUDY00002661). Additionally, we will obtain consent from all involved participants before collecting implementation data. In the implementation phase, organizations will receive community-informed implementation support focusing on intervention participants (e.g., providing transportation) and/or focusing on organizations (e.g., cultural competency training) based on information gathered in the exploration phase of the project and developed and tailored in the preparation phase. The overarching goal of SFF is to reduce behavioral health disparities and improve family resilience in the Flint Community.

The RE-AIM model will be used to guide the implementation evaluation. Designed to evaluate public health interventions, RE-AIM focuses on the assessment of five dimensions (i.e., Reach, Effectiveness, Adoption, Implementation, Maintenance); these dimensions will be assessed at the multiple levels of our research initiative (i.e. individual, family, and community) (Glasgow et al., 2019). The multi-level nature of RE-AIM provides a natural alignment with the design of SFF.

### Aim 1: Create a network of organizations that taps into the natural strengths of the community and is committed to Strengthening Flint Families

To accomplish our first goal of strengthening connections between existing organizations to effectively leverage existing services in the Flint Community, we first sought to better understand the services currently offered within the community. Second, we connected individual service providers to form a network of organizations serving the behavioral health needs of the Flint community. We accomplished these goals by: 1) identifying existing behavioral health organizations and conducting key informant interviews to validate the listings, and identify non-licensed providers and other potential stakeholders; 2) conducting organizational assessments to identify what existing behavioral health prevention, treatment and recovery services are available; and 3) creating formal networking opportunities to align and streamline these existing resources.

#### Identifying Organizations for Inclusion

The preparation phase began by identifying organizations in Flint with a stake in behavioral health. The Michigan Department of Licensing and Regulatory Affairs Bureau of Health Systems’ License Listing Report contains all behavioral health sites licensed to provide direct services (including inpatient and outpatient drug treatment programs, mental health centers, etc.) within Genesee County that provide some type of behavioral health service. Seventeen of these sites are within the City of Flint and were identified as potential sites. One of these sites, Flint Odyssey House, was already represented as the community partner for this research project. In addition to licensed behavioral health providers, many community organizations, coalitions, and other grass-roots organizations facilitate behavioral health and health equity work in Flint. The Flint Neighborhoods United and The Flint Water Recovery Group databases were used to identify forty-two community-based organizations with missions related to behavioral health. Schools typically host programming for families and adolescents and are often a convenient location for families (National Research Council, 2000; Johnstone, Kemps, & Chen, 2018). Eleven schools were identified through flintschools.org as potential sites. Additionally, churches are in a unique position to serve as facilitators of and referrers to behavioral health services in their communities (Chalfant et al., 1990; Florez et al., 2019), but were not included at this time because of their central role in the FCHES other major project, (the Church Challenge) (Johnson-Lawrence et al., 2019).

All organizations were considered as potential contributors of a synergistic community network of adopters, hosts, and referrers. These categories are not mutually exclusive. Adopters are organizations that will be able to directly deliver SFF services. These organizations will host, staff, and provide PRCs and/or SFP sessions to the population they serve. Hosts are organizations that have the physical space and accessibility requirements, but do not have the ability to directly deliver (i.e., adopt) the program due to a lack of staff or appropriate credentials. Referrers (e.g., Parole and Probation, WIC, etc.) are organizations within the community that regularly interact with the focus population and provide resources and/or assistance programs. Using this framework, we assessed the interests and capacities of each organization to determine how they could best operate within this network.

### Aim 2: Tailor implementation strategies for peer recovery coaching, the Strengthening Families Program, and a multi-media health promotion campaign and add them to services

To best understand the needs of the community and to inform the development of implementation strategies, we will use data collected from key informant interviews and organization assessments.

#### Key Informant Interviews

Key informant interviews were conducted with 9 community stakeholders identified by FCHES members and the project leadership team representing different aspects of the community that are crucial to the success of the SFF initiative. Community experts in behavioral health treatment and prevention programs, family and youth programs, and the faith community were all represented. The semi-structured interviews included questions about barriers that individuals, families, and organizations face when engaging with treatment or prevention programs, and other local knowledge that could affect participation or community engagement. Feedback on the list of potential sites was also requested, especially for organizations in that expert’s field.

#### Organizational Assessments

The research team developed organizational assessments to assess each organization’s interest in collaborating, to learn relevant information about each organization, and to identify potential organizational collaborators. These assessments also doubled as opportunities to generate enthusiasm around this behavioral health initiative. The research team contacted each site, identified through the process described in the above section, and administered the assessment to a mid-to-high level organization leader. The ideal organization leader would have direct experience with that organization’s day-to-day activities while also having enough authority to make decisions about the initiatives of their organization or unit within a larger organization. At the end of the assessment, each organization leader was asked if they knew of any other organizations in the Flint area that they thought would be interested in the SFP or PRC. These suggestions were recorded by the research team and added to the list of potential organizations if they met the criteria of the original list. This was done to ensure that no organizations were left out of this initiative.

### Aim 3: Build capacity and promote sustainability

An objective of our implementation protocol is to increase connections between existing organizations and services and formally develop the SFF network. The network is designed to increase the number of referrals between organizations, the appropriateness of referrals to services, reduce program overlap, and support navigations of referrals to actual services. All organizations who participated in the organization assessment will be provided with referral information and encouraged to refer individuals and families to the SFP or PRC services. Additionally, we will offer continuing education programming and other networking opportunities for PRCs. Networking and programming opportunities will be available throughout the implementation period and will allow for both formal and informal opportunities to interact with other providers and organizations serving the Flint community. Second, we will work with a local non-profit agency, the Greater Flint Health Coalition, to expand their existing electronic referral system, a clearinghouse of behavioral and physical health care providers in the Flint community. We will use the results of the key informant interviews and organizational assessments to identify agencies that can be added to the referral system, and will promote the system among existing providers during our networking events, in order to increase interconnections between agencies.

In the implementation phase, behavioral health organizations, churches, schools, and other community organizations will be identified as adopters, hosts, and referrers based on the organizational assessments, and will be invited to enroll in a synergistic community network. This strategy was identified to improve the reach and sustainability of SFF. Reach will increase as a result of the availability of multiple host and/or adopter sites located in different areas of the city, placing programs closer to community members to reduce the burden and increase access (Syed, Gerber, & Sharp, 2013).

Sustainability will be facilitated by carefully selecting the number, geographic location, and appropriateness of organizations as adopters. A systematic review of public health evidence-based interventions in community settings found that lack of funding independent of research grants is the most common barrier to sustainability (Hailemariam et al., 2019). Organizations with ample funding and infrastructure to consistently offer and develop programs with existing revenue streams, independent of research dollars, will be considered for potential adopter sites. Adopter sites are also encouraged to have a strong connection to the community to generate a sustainable referral base, which will be strengthened by the SFF network. These few adopter sites can then offer programming at one of the many host sites, therefore optimizing sustainability and reach in a scalable fashion.

In the implementation phase, all organizations adopting or hosting SFP or PRCs will receive community-informed implementation support focusing on intervention participants (e.g., providing transportation) and/or focusing on organizations (e.g., cultural competency training) based on information gathered in the key informant interviews and continued evaluation of the success of these strategies. The recommended strategies will reflect the barriers that their specific population and/or organization faces.

### Aim 4: Evaluate primary (implementation) and secondary (effectiveness) outcomes

#### Implementation Outcomes

Multiple sources of data will be used to synthesize our findings within the RE-AIM framework and describe the cumulative impact of this community-partnered effort to promote behavioral health. *Reach*, or the proportion of the eligible population that participates in the intervention (Glasgow et al., 2019), will be assessed by documenting the number of eligible participants, the number/percent of those eligible consenting to participate and the percent of participants retained during the study period. Described in greater detail below, we will examine *effectiveness* of the *SFP 10-14* by examining changes in program participants pre- and post-intervention, as well as community-level changes in substance use and associated problems.

*Adoption* refers to the representativeness of settings in which the intervention takes place (Glasgow et al., 2019). We will assess facilitators of, and barriers to, adoption through interviews with program staff and administrators. Willingness of organizations to participate (whether adopting, hosting, or referring) in the *SFF* framework will be evaluated as a measure of adoption at the organization level. Within organizations, staff will be surveyed on their willingness to participate in the programs. Unique themes will be identified in an inductive manner through iterative stages. Narrative text will be reviewed independently by two researchers to identify themes and sub-themes focused on facilitators of, and barriers to, adoption. Emergent themes will be logged, defined, and sample text illustrating the theme will be noted. Using constant comparative technique, themes will be repeatedly reviewed to determine the need to expand or merge thematic codes. The analysis will be conducted manually. The researchers will compare findings and, if needed, achieve consensus through discussion. Surveys of organization staff on their willingness to use the programs, at the beginning and end of program funding, will be analyzed using a paired samples *t*-test.

*Implementation* describes whether the intervention is delivered as designed (i.e., fidelity, acceptability, and feasibility) (Glasgow et al., 2019). Using established implementation quality tools developed by the SFP creators (Spoth et al., 2014), we will assess the (a) degree of adherence to the curriculum, (b) participant interest and engagement, and (c) facilitator effectiveness. SFP facilitators will consist of local organization staff and will 1) maintain attendance records of participants; and 2) maintain SFP session logs (e.g. curriculum activities completed or skipped). Research staff will conduct systematic on-site observations of curriculum sessions. Select sessions will be observed for fidelity. Using a standard observation format created by the SFP (Spoth et al, 2014, Kumpfer & Brown, 2017), research staff will record completion of curriculum activities, engagement of youth and parents, and external factors that may affect participation. Acceptability will be assessed with standard SFP brief program evaluation/satisfaction surveys at the end of each cohort. Attendance data, session logs, and program evaluation/satisfaction data will be de-identified and electronically transferred to the research team at the end of each SFP series.

Peer recovery coaching activities are highly individualized based on the needs of the client. PRCs develop a plan with their client that aligns with domains important to recovery (e.g., employment) and traditional measures of fidelity are not applicable to this model of service delivery. Instead, acceptability and feasibility will be determined using focus groups with PRCs that focus on their experiences implementing recovery coaching in the Flint population. Specifically, PRCs will be asked to report the barriers that they have encountered when working with individuals in recovery, with a specific focus on how feasible it is to deliver services using this model and how they perceive clients’ engagement in their services. We will transcribe the recordings from the PRC focus group to use as qualitative data. All de-identified information will be removed from the transcripts. Common themes will be identified from the focus group. The themes will be utilized to guide our work in building a strong network of local PRCs and connecting PRCs to community-serving organizations.

*Maintenance* describes the degree to which an intervention becomes an integral part of an institutional structure (Glasgow et al., 2019). Evaluation and feedback are critical components of the sustainability, or maintenance, phase of evidence-based practices (Schoenwald et al., 2011). We will document how many of the participating organizations (adoption sites) implement the program in the year following the completion of implementation. We will also measure the maintenance of our referral network by analyzing data on referrals over time.

#### Effectiveness Outcomes

Although the primary goal of this study is to assess implementation, a secondary goal is to assess and monitor the effectiveness of the programs (i.e., the SFP, and the multi-media campaign).

For the family-level SFP intervention, we will obtain pre-post survey data using the standard SFP surveys (available through the Evidence-Based Prevention & Intervention Support Center, http://epis.psu.edu/ebp/strengthening/outcomemeasurementtools) from parents and youth upon entry into the program and upon completion of the 7-week intervention from the organizations. The survey focuses on assessing the outcomes of family functioning and communication. Data will be de-identified and electronically transferred to the research team for analysis. For analysis, we will link participant pre-post test data to their family, cohort, facilitator, and organization. Paired T tests will be conducted to evaluate differences in outcomes before and after the program. We will also analyze the data to evaluate any possible differential effects by the various grouping variables (e.g. organization, family, cohort, or facilitators).

Community-level effectiveness outcomes of this project include substance use and substance use treatment data. Substance use data will be obtained through a community-wide survey of Flint residents called the *Speak to Your Health Survey*. The *Speak to Your Health Survey* is a biennial survey of Flint residents that began in 2003 to monitor the individual and community-level impacts of public health initiatives. Substance use treatment data will be obtained from insurance data from the Genesee Health Plan. Additionally, we will track how organizations are prompted to generate a family referral to the SFP to better understand the best mechanisms for promoting referrals to the program, and the program itself. We will also track social media analytics to assess the level of engagement our social media pages have with the community.

## Discussion

Previous studies have documented the need for supplementing effectiveness research with research designs that specifically examine the effect of different implementation conditions on the reach, effectiveness, adoption, implementation and sustainment of interventions (Chaudoir, Dugan, & Barr, 2013). We expect that the evidence produced as a result of this work will contribute towards a better understanding of the importance of purposeful and systematic plans to facilitate reach, successful adoption and better sustainment of behavioral health programs in cities with high levels of inequity. Furthermore, we expect to learn a great deal about the relative effectiveness of different implementation strategies in low-resource settings.

## Data Availability

This manuscript contains no data from individuals for publication.

## List of Abbreviations

SFF: Strengthening Flint Families
FCHES: Flint Center for Health Equity Solutions
PRC: Peer Recovery Coach
SFP: Strengthening Families Program
CBPR: Community-Based Participatory Research
RE-AIM: Reach, Effectiveness, Adoption, Implementation, Maintenance

## Declarations

## Ethics approval and consent to participate

This study was approved by the Michigan State University Institutional Review Board on May 28, 2019 and was “determined not to involve ‘human subjects’ as defined by the Common Rule as codified in the U.S. Department of Health and Human Services (DHHS) regulations for the protection of human research subjects”.

## Consent for publication

This manuscript contains no data from individuals for publication.

## Authors’ contributions

SS, DFH, KSY, JF, BWM, LDM, AD, and JJ made substantial contributions to the conception and design of this work.

BWM, SS, JF, DFH, and LDM have drafted the work or substantively revised it.

SS, DFH, KSY, JF, BWM, LDM, AD, and JJ reviewed and approved the final manuscript.

## Acknowledgements

The authors wish to thank Richard Sadler, Ph.D. for his contribution of material on the study setting and his methodology for choosing sites to be assessed. The authors wish to thank the community organizations and key informants who participated in the initial assessments and network development.

## References

1. Barclay C, Viswanathan M, Ratner S, Tompkins J, Jonas DE. Implementing evidence-based screening and counseling for unhealthy alcohol use with epic-based electronic health record tools. The Joint Commission Journal on Quality and Patient Safety. 2019;45(8):566–74.

2. Bernet AC, Willens DE, & Bauer MS. Effectiveness-implementation hybrid designs: implications for quality improvement science. Implementation Science: IS, 2013;8(Suppl 1), S2. http://doi.org/10.1186/1748-5908-8-S1-S2.

3. Boisvert RA, Martin LM, Grosek M, Clarie AJ. Effectiveness of a peer-support community in addiction recovery: participation as intervention. Occupational Therapy International. 2008;15(4):205–20.

4. Chalfant HP, Heller PL, Roberts A, Briones D, Aguirre-Hochbaum S, & Farr W. The clergy as a resource for those encountering psychological distress. Review of Religious Research, 1990;305–313.

5. Chaudoir SR, Dugan AG, & Barr CH. Measuring factors affecting implementation of health innovations: a systematic review of structural, organizational, provider, patient, and innovation level measures. Implementation Science, 2013;8(1), 22.

6. Chinman M, George P, Dougherty RH, Daniels AS, Ghose SS, Swift A, Delphin-Rittmon ME. Peer support services for individuals with serious mental illnesses: assessing the evidence. psychiatric services. 2014 Apr 1;65(4):429–41. Doi: 10.1176/appi.ps.201300244.

7. DeMarsh J, & Kumpfer KL. Family-oriented interventions for the prevention of chemical dependency in children and adolescents. Journal of Children in Contemporary Society, 1986;18(1-2), 117–151.

8. Dugosh K, Abraham A, Seymour B, McLoyd K, Chalk M, Festinger D. A systematic review on the use of psychosocial interventions in conjunction with medications for the treatment of opioid addiction. J Addict Med. 2016;10(2):93–103. doi:10.1097/ADM.0000000000000193.

9. Ellington R, Scott JB, Barajas CB, Meghea C, Drahota A, Uphold H, Furr-Holden CDM. An evaluation framework of a transdisciplinary collaborative center for health equity research. American Journal of Evaluation. Forthcoming 2020.

10. Florez KR, Payan DD, Palar K, Williams MV, Katic B, Derose KP. Church-based interventions to address obesity among African Americans and Latinos in the United States: a systemic review. Nutrition Reviews. 2019;78(4):304–322.

11. Glasgow RE, Harden SM, Gaglio B, Rabin B, Smith ML, Porter GC, Ory MG, Estabrooks PA. RE-AIM planning and evaluation framework: adapting to new science and practice with a 20-year review. Frontiers in Public Health. 2019 Mar 29;7:64.

12. Gottfredson D, Kumpfer K, Polizzi-Fox D, Wilson D, Puryear V, Beatty P, & Vilmenay M. The strengthening Washington DC families project: a randomized effectiveness trial of family-based prevention. Prevention Science, 2006;7(1), 57–74.

13. Green LW, George MA, Daniel M, et al. Study of participatory research in health promotion: review and recommendations for the development of participatory research in health promotion in Canada. Vancouver, British Columbia: Royal Society of Canada; 1995:4.

14. Green LW, Glasgow RE, Atkins D, Stange K. Making evidence from research more relevant, useful, and actionable in policy, program planning, and practice: slips “twixt cup and lip”. American Journal of Preventive Medicine, 2009;37(6):S187–S191.

15. Griffith DM, Allen JO, DeLoney EH, Robinson K, Lewis EY, Campbell B, Morrel-Samuels S, Sparks A, Zimmerman MA, Reischl T. Community-based organizational capacity building as a strategy to reduce racial health disparities. The Journal of Primary Prevention, 2010;31(1-2), 31–39.

16. Hailemariam M, Bustos T, Montgomery B, Barajas R, Evans L, Drahota A. Evidence-based intervention sustainability strategies: a systematic review. IS, 2019;12. https://doi.org/10.1186/s13012-019-0910-6.

17. Hanna-Attisha M, LaChance J, Sadler RC, and Schnepp AC. Elevated blood lead levels in children associated with the flint drinking water crisis: a spatial analysis of risk and public health response. American Journal of Public Health, 2015;106(2), 283–290.

18. Israel BA, Schulz AJ, Parker EA, Becker AB, Allen AJ, Guzman JR, & Lichtenstein R. Critical issues in developing and following CBPR principles. In: Wallerstein N, Duran B, Oetzel J, Minkler M, editors. Community-Based participatory research for health: advancing social and health equity. 2008. P. 31.

19. Jacobs AJ. The impacts of variations in development context on employment growth: a comparison of central cities in Michigan and Ontario, 1980-2006. Economic Development Quarterly, 2009;23(4), 351–371.

20. Johnson-Lawrence V, Bailey S, Sanders PE, Sneed R, Angel-Vincent A, Brewer A, et al. The church challenge: a community-based multilevel cluster randomized controlled trial to improve blood pressure and wellness in African American churches in Flint, Michigan. Contemporary Clinical Trials Communications. 2019;14:100329.

21. Johnstone KM, Kemps E, Chen J. A meta-analysis of universal school-based prevention programs for anxiety and depression in children. Clinical Child and Family Psychology Review. 2018 Aug 14;21(4):466–81.

22. Kilbourne AM, Abraham KM, Goodrich DE, Bowersox NW, Almirall D, Lai Z, Nord KM. Cluster randomized adaptive implementation trial comparing a standard versus enhanced implementation intervention to improve uptake of an effective re-engagement program for patients with serious mental illness. Implementation Science, 2013 Nov 20;8(1). http://doi.org/10.1186/1748-5908-8-136.

23. Kumpfer KL, & DeMarsh J. Family environmental and genetic influences on children’s future chemical dependency. Journal of Children in Contemporary Society, 1986;18(1-2), 49–91.

24. Kumpfer KL, Alvarado R, Smith P, & Bellamy N. Cultural sensitivity and adaptation in family-based prevention interventions. Prevention Science, 2002;3(3), 241–246.

25. Kumpfer K, Brown J. SFP 7-17 process fidelity observation & family coach self-rating forms [measurement instrument]. Retrieved from authors. 2017.

26. Liebling EJ, Yedinak JL, Green TC, Hadland SE, Clark MA, Marshall BDL. Access to substance use treatment among young adults who use prescription opioids non-medically. Substance Abuse Treatment, Prevention, and Policy. 2016;11(1).

27. MacArthur GJ, Harrison S, Caldwell DM, Hickman M, and Campbell R. PeerJled interventions to prevent tobacco, alcohol and/or drug use among young people aged 11– 21 years: a systematic review and metaJanalysis. Addiction, 2016;111:391–407. doi:10.1111/add.13224.

28. National Research Council. After-School Programs that Promote Child and Adolescent Development: Summary of a Workshop. National Academies Press. 2000.

29. Robinson MN, Tansil KA, Elder RW, Soler RE, Labre MP, Mercer SL, et al. Mass media health communication campaigns combined with health-related product distribution. American Journal of Preventive Medicine. 2014 Sep;47(3):360–71.

30. Romero LM, Olaiya O, Hallum-Montes R, Varanasi B, Mueller T, House LD, et al. Efforts to increase implementation of evidence-based clinical practices to improve adolescent-friendly reproductive health services. Journal of Adolescent Health. 2017;60(3).

31. Sadler RC, Gilliland JA, Arku G. A food retail-based intervention on food security and consumption. International Journal of Environmental Research and Public Health, 2013;10(8), 3325–3346.

32. Sadler RC, Highsmith AR. Rethinking tiebout: the contribution of political fragmentation and racial/economic segregation to the Flint water crisis. Environmental Justice, 2016;9(5), 143–151.

33. Sadler RC, Lachance J, Hanna-Attisha M. Social and built environmental correlates of predicted blood lead levels in the Flint water crisis. American Journal of Public Health. 2017;107(5):763–9.

34. Sadler RC, Furr-Holden D. The epidemiology of opioid overdose in Flint and Genesee County, Michigan: implications for public health practice and intervention. Drug and Alcohol Dependence. 2019 Nov;204:107560. doi: 10.1016/j.drugalcdep.2019.107560. [Epub ahead of print].

35. Schoenwald SK, Garland AF, Chapman JE, et al. Adm Policy Ment Health. 2011;38:32. https://doi.org/10.1007/s10488-010-0321-0.

36. Shaver ER, Sadler RC, Hill AB, Bell K, Ray M, Choy-Shin J, Lerner J, Soldner T, Jones AD. The Flint food store survey: combining spatial analysis with a modified Nutrition Environment Measures Survey in Stores (NEMS-S) to measure the community and consumer nutrition environments. Public Health Nutrition, 2018;21(8), 1474–1485.

37. Speak To Your Health! Community Survey Results Tables. In: Speak To Your Health. Genesee County Health Department; 2014. http://speak.gchd.us/wp-content/uploads/2016/10/2013-Community-Survey-Results-Tables.pdf. Access 25 Nov 2019.

38. Spoth R, Redmond C, Shin C, & Azevedo K. Brief family intervention effects on adolescent substance initiation: school-level growth curve analyses 6 years following baseline. Journal of Consulting and Clinical Psychology, 2004;72(3), 535.

39. Spoth R, Redmond C, & Shin C. Direct and indirect latent-variable parenting outcomes of two universal family-focused preventive interventions: extending a public health-oriented research base. Journal of Consulting and Clinical Psychology, 1998;66(2), 385.

40. Spoth R, Trudeau L, Redmond C, Shin C. Replication RCT of early universal prevention effects on young adult substance misuse. Journal of Consulting and Clinical Psychology. 2014;82(6):949–63.

41. Substance Abuse and Mental Health Services Administration. (2017a). Key substance use and mental health indicators in the United States: results from the 2016 National Survey on Drug Use and Health (HHS Publication No. SMA 17-5044, NSDUH Series H-52). Rockville, MD: Center for Behavioral Health Statistics and Quality, Substance Abuse and Mental Health Services Administration. https://www.Samhsa.gov/data/.

42. Substance Abuse and Mental Health Services Administration. (2017b). Peers supporting recovery from substance use disorders. In: Substance Abuse and Mental Health Services Administration.https://www.samhsa.gov/sites/default/files/programs_campaigns/brss_tacs/peers-supporting-recovery-substance-use-disorders-2017.pdf. Accessed 25 Nov 2019.

43. Syed ST, Gerber BS, & Sharp LK. Traveling towards disease: transportation barriers to health care access. Journal of Community Health, 2013;38(5), 976–993.

44. US Census Bureau. In: US Census Bureau. 2017. https://www.census.gov/.

45. Wang PS, Lane M, Olfson M, Pincus HA, Wells KB, Kessler RC. Twelve-month use of mental health services in the United States: results from the National Comorbidity Survey Replication. Arch Gen Psychiatry. 2005;62(6):629–640. doi:10.1001/archpsyc.62.6.629.

46. Wen H, Druss BG, & Cummings JR. Effect of Medicaid expansions on health insurance coverage and access to care among lowJincome adults with behavioral health conditions. Health Services Research, 2015;50(6), 1787–1809. http://doi.org/10.1111/1475-6773.12411

